# Circulating levels of calcitonin gene-related peptide (CGRP) are lower in COVID-19 patients

**DOI:** 10.1101/2020.10.01.20205088

**Authors:** Laura Ochoa-Callejero, Josune García-Sanmartín, Pablo Villoslada-Blanco, María Íñiguez, Patricia Pérez-Matute, Rachel Brody, José A. Oteo, Alfredo Martínez

**Author notes:** Correspondence to: Alfredo Martínez, Angiogenesis Group, Oncology Area, Center for Biomedical Research of La Rioja (CIBIR), Piqueras 98, 26006 Logroño, Spain. Conflict of interests: Authors declare that no conflict of interest exists.

## Abstract

**Background:** To better understand the biology of COVID-19, we have explored the behavior of calcitonin gene-related peptide (CGRP), an angiogenic, vasodilating, and immune modulating peptide, in SARS-CoV-2 positive patients.

**Methods:** Levels of CGRP in the serum of 57 COVID-19 patients (24 asymptomatic, 23 hospitalized in the general ward, and 10 admitted to the intensive care unit) and healthy donors (n=24) were measured by ELISA. In addition, to better understand the physiological consequences of the observed variations, we investigated by immunofluorescence the distribution of RAMP1, one of the components of the CGRP receptor, in autopsy lung specimens.

**Results:** CGRP levels greatly decreased in COVID-19 patients (p<0.001) when compared to controls, and there were no significant differences due to disease severity, sex, age, or comorbidities. We found that COVID-19 patients treated with proton pump inhibitors had lower levels of CGRP than other patients not taking this treatment (p=0.001). RAMP1 immunoreactivity was found in smooth muscle cells of large blood vessels and the bronchial tree, and in the airways epithelium. In COVID-19 samples, RAMP1 was also found in proliferating type II pneumocytes, a common finding in these patients.

**Conclusions:** The lower levels of CGRP should negatively impact the respiratory physiology of COVID-19 patients due to vasoconstriction, improper angiogenesis, less epithelial repair, and faulty immune response. Therefore, restoring CGRP levels in these patients may represent a novel therapeutic approach for COVID-19.

## Introduction

The COVID-19 pandemic has developed into the major challenge to global health in our lifetime. This disease is caused by severe acute respiratory syndrome coronavirus 2 (SARS-CoV-2) and it affects people in different ways, ranging from asymptomatic disease to severe pneumonia than may require oxygen therapy, and even leading to death ^1^. At present there is no specific treatment for the disease and only prevention strategies such as promoting social distance, increasing general hygiene, and applying symptomatic remedies can reduce COVID-19 impact ^2^. Also, vaccine development is progressing rapidly ^3^. A full understanding of the biology implicated in the evolution of the disease may provide new ideas for the treatment and management of patients. In this regard, interesting data are arising from the study of autopsy specimens ^4^. Among other pathological findings, the occurrence of angiogenesis in the lung of seriously affected patients has been reported ^5^. A recent article has studied the potential pro-angiogenic molecules whose expression is upregulated in COVID-19 patients ^6^. Among those markers, we were especially interested in receptor activity modifying protein 1 (RAMP1) which, in combination with the calcitonin receptor-like receptor (CLR), constitutes the receptor for calcitonin gene-related peptide (CGRP) ^7^.

CGRP is a pro-angiogenic molecule ^8^ but it also plays other important roles in the lung including vasoregulation, bronchoprotection, anti-inflammatory actions, and tissue repair ^9, 10^, all of them very relevant for the evolution of COVID-19 pathogenesis. In the normal lung, CGRP is found in neuroepithelial bodies and in nerve fibers which contact the epithelium, neuroendocrine cells, and smooth muscle ^11^. CGRP binding sites have been found in pulmonary blood vessels, and the smooth muscle and epithelium of large airways ^12^. It has been shown that CGRP promotes the growth of bronchial ^13^ and alveolar ^14^ epithelial cells following lung injury, thus acting as a protective mediator in all kinds of experimental procedures involving lung insults ^15^.

Another important function for CGRP in the lung is the regulation of the immune system. CGRP expression is enhanced in response to inflammation ^16^ and it exerts anti-inflammatory actions by regulating macrophage polarization ^17^ and by inhibiting dendritic and T cells ^18^. CGRP also limits group 2 innate lymphoid cell (ILC2) responses ^19^. Depending on the context, this anti-inflammatory potential may be positive or negative for the patient. For instance, the presence of CGRP-positive nerves reduced immunity and thus survival in a model of lethal *Staphylococcus aureus* pneumonia ^20^. In addition, in knock-out models, where either the peptide ^21^ or the receptor ^22^ were disrupted, there was a reduction of allergic asthma responses. This involvement with the immune system could be critical for the pathophysiology of COVID-19 ^23^.

Therefore, our objective was to evaluate the levels of circulating CGRP in COVID-19 patients with different symptoms and to compare them with healthy controls. We also studied the expression of RAMP1 in the lungs of patients who died of COVID-19 and compared them with patients who died by unrelated causes.

## Methods

### Serum samples

Blood samples were obtained at Hospital San Pedro (Logroño, Spain) from healthy volunteers recruited before the initiation of the pandemia (n=24), and from COVID-19-positive patients (confirmed by PCR), which were subdivided in 3 groups depending on disease severity: i) asymptomatic or mildly symptomatic patients not requiring hospitalization (n=24), ii) patients requiring hospitalization in the normal ward (n=23), and iii) patients admitted to the intensive care unit (ICU) (n=10). Blood was collected in serum separator tubes (BD Vacutainer, Becton Dickinson, Franklin Lakes, NJ) and serum was isolated, aliquotted, and frozen at −80°C until further analysis.

Relevant clinical data were obtained from the clinical history of the patients. All procedures were approved by the local review board (Comité de Ética de Investigación con Medicamentos de La Rioja, CEImLAR, ref. PI-412). All described procedures adhere to the tenets of the Declaration of Helsinki.

### Autopsy specimens

Paraffin tissue sections from the lung of three patients who died from COVID-19 and from three other patients who died from pathologies unrelated to COVID-19, and with no lung involvement, were generated at the Department of Pathology of the Icahn School of Medicine at Mount Sinai (New York), and sent to Spain for analysis.

### CGRP ELISA protocol

Levels of CGRP were quantitated in serum samples using a commercial enzyme-linked immunosorbent assay (ELISA) kit (MBS2023906, MyBioSource, San Diego, CA), following manufacturer’s instructions.

### Immunofluorescence and confocal microscopy

Tissue sections were dewaxed, rehydrated, and subjected to antigen retrieval (10mM Sodium Citrate, 0.5% Tween 20, pH 6.0, 20 min at 95°C). Non-specific binding was blocked by exposure to 10% normal donkey serum (Jackson Immunoresearch Laboratories, West Grove, PA) for 1 h, and then tissue sections were incubated with recombinant rabbit monoclonal anti-RAMP1 antibody, clone EPR10867 (ab156575, Abcam), overnight at 4°C. The following day, sections were incubated with fluorescent secondary antibody, CF633 donkey anti-rabbit IgG (20125, Biotium, Fremont, CA), for 1 h. Finally, they were counterstained with DAPI (Molecular Probes, Eugene, OR) and analyzed with a confocal microscope (TCS SP5, Leica, Badalona, Spain). Negative controls were performed by substituting the primary antibody by PBS.

### Statistical analysis

All data were analyzed with GraphPad Prism 8 software and were considered statistically significant when p<0.05. Values are expressed as means ± SEM. Normally distributed data were evaluated by Student’s *t* test or by ANOVA followed by the Dunnet’s post-hoc test, while data not following a normal distribution were analyzed with the Kruskal-Wallis test followed by the Mann-Whitney U test. Categorical data were analyzed with Fisher’s exact test.

## Results

Serum samples were collected from healthy volunteers (n=24) and from PCR-confirmed COVID-19 patients (n=57) whose disease had different degrees of severity: asymptomatic (n=24), hospitalized in the general ward (n=23), or admitted to the intensive care unit (ICU) (n=10). Three of the patients in the ICU group eventually died of COVID-19 complications. Age distribution of the healthy control population was lower than that of the COVID-19 group but no differences were found in gender distribution (Table 1). As expected, patients with more serious disease were older (p<0.001) and had a higher number of comorbidities, including hypertension (p<0.001) and dyslipidemia (p=0.02). They were also more likely to be taking chronic medication such as angiotensin-converting enzyme (ACE) inhibitors (p=0.007), antidiuretics (p=0.007), antidepressants (p=0.007), or proton pump inhibitors (p=0.008) (Table 2). CGRP levels were measured in the serum of all subjects by ELISA. Healthy volunteers’ CGRP levels were 289.9 ± 35.9 pg/ml, whereas COVID-19 patients presented significantly lower levels: 105.7 ± 7.0 pg/ml (p<0.001). When separated by disease severity, the three COVID-19 groups displayed significantly lower levels of CGRP when compared to healthy controls, but there were no significant differences among disease groups (Fig. 1A). No differences were found in CGRP levels by sex or age in either the healthy or the disease groups (results not shown). When testing comorbidities and chronic drug treatments, we found that patients receiving proton pump inhibitors had significantly lower levels of CGRP than the rest of COVID-19 patients (p=0.001) (Fig. 1B). To test whether these pump inhibitors were responsible for the total reduction in CGRP levels observed in the COVID-19 population, we reanalyzed the data after removing all patients treated with the inhibitors (n=12). The remaining patients (n=45) had CGRP levels of 110.2 ± 7.6 pg/ml, and were still significantly lower than the healthy control levels (p<0.001). All other conditions did not significantly affect CGRP levels.

**Table 1.** Baseline characteristics of the study population

|  | **Controls** | **COVID-19** | ***p*** |
| --- | --- | --- | --- |
| *n* | 24 | 57 |  |
| Age, mean ± SEM | 44.3 ± 2.4 | 58.3 ± 2.5 | **0.002 ¶** |
| Sex (Male) | 9 (37.5 %) | 25 (43.9 %) | 0.63 † |
| CGRP (pg/ml), mean ± SEM | 289.9 ± 35.9 | 105.7 ± 7.0 | **<0.001 ¶** |

¶: U Mann-Whitney ; †: Fisher. Significant differences appear in bold case.

**Table 2.** Clinical characteristics of the COVID-19 patients included in the study

|  |  | Asymptomatic | Hospital ward | ICU | *P* * |
| --- | --- | --- | --- | --- | --- |
| **n** |  | 24 | 23 | 10 |  |
| **Age, mean ± SEM** | | 44.3 ± 1.9 | 72.3 ± 4.1 | 59.9 ± 2.8 | **<0.001 ¶** |
| **Sex (Male)** |  | 8 (33.3 %) | 9 (39.1 %) | 8 (80 %) | 0.19 † |
| **Risks factors** |  |  |  |  |  |
|  | Arterial hypertension | 0 (0 %) | 13 (56.5 %) | 4 (40 %) | **<0.001 †** |
|  | Dyslipidemia | 0 (0 %) | 6 (26.1 %) | 2 (20 %) | **0.02 †** |
|  | Diabetes | 0 (0 %) | 4 (17.4 %) | 0 (0 %) | 0.13 † |
|  | Obesity | 0 (0 %) | 2 (8.7 %) | 2 (20 %) | 0.13 † |
|  | Ischemic cardiopathy | 0 (0 %) | 3 (13.0 %) | 1 (10 %) | 0.13 † |
|  | Arrhythmia | 0 (0 %) | 3 (13.0 %) | 1 (10 %) | 0.13 † |
|  | EPOC | 0 (0 %) | 4 (17.4 %) | 0 (0 %) | 0.13 † |
| **Previous treatment** |  |  |  |  |  |
|  | ACE inhibitors | 0 (0 %) | 6 (26.1 %) | 3 (30 %) | **0.007 †** |
|  | Statins | 1 (4.2 %) | 5 (21.7 %) | 2 (20 %) | 0.12 † |
|  | Antidiuretics | 0 (0 %) | 7 (30.4 %) | 2 (20 %) | **0.007 †** |
|  | Antidepressants | 0 (0 %) | 8 (34.8 %) | 1 (10 %) | **0.007 †** |
|  | H<sup>+</sup> pump inhibitors | 1 (4.2 %) | 9 (39.1 %) | 2 (20 %) | **0.008 †** |
|  | Beta blockers | 1 (4.2 %) | 3 (13.0 %) | 3 (30 %) | 0.22 † |
|  | Anticoagulants | 0 (0 %) | 4 (17.4 %) | 0 (0 %) | 0.13 † |
| **CGRP (pg/ml), mean ± SEM** | | 100.9 ± 7.6 | 97.6 ± 10.7 | 135.9 ± 24.9 | 0.80 ¶ |

\* = p value comparing mild (Asymptomatic) vs severe (Hospital Ward and ICU) disease cases.
¶: U Mann-Whitney ; †: Fisher. Significant differences appear in bold case.

**Figure 1.**
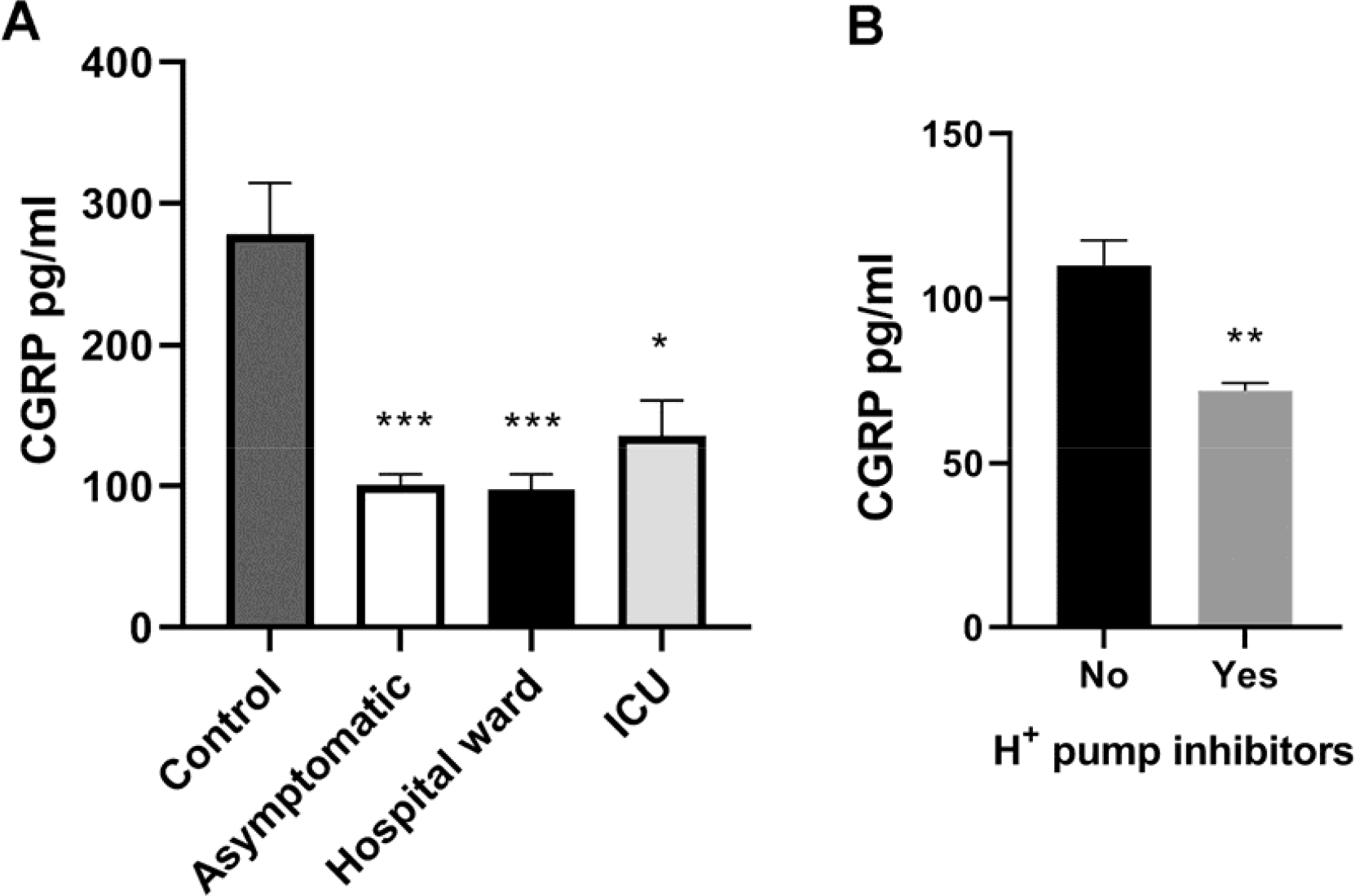
Serum CGRP levels as measured by ELISA. All COVID-19 positive patients (asymptomatic, hospital ward, or ICU) had significantly lower CGRP levels than healthy controls (A). Bars represent mean ± standard error of the mean. Patients receiving chronic treatment with proton pump inhibitors had significantly lower CGRP levels than patients not receiving them (B). *: p<0.05; **: p<0.01; ***: p<0.001 compared to control.

In order to better understand the physiological consequences in the lung of the systemic reduction in CGRP levels, we investigated by immunofluorescence the distribution of the CGRP receptor component RAMP1. RAMP1 immunoreactivity was studied in lung sections of three patients who died from COVID-19 complications and of three other patients who died from causes unrelated to COVID-19, with no lung pathology. In the normal lung, RAMP1 immunoreactivity was predominantly found in the muscle layer surrounding large blood vessels (Fig. 2A) and major airways, and, with lower intensity, in the bronchial and bronchiolar epithelium (Fig. 2C). No RAMP1 immunoreactivity was found in the alveolar epithelium. Absence of the primary antibody completely precluded labeling, thus confirming staining specificity (Fig. 2B,D).

**Figure 2.**
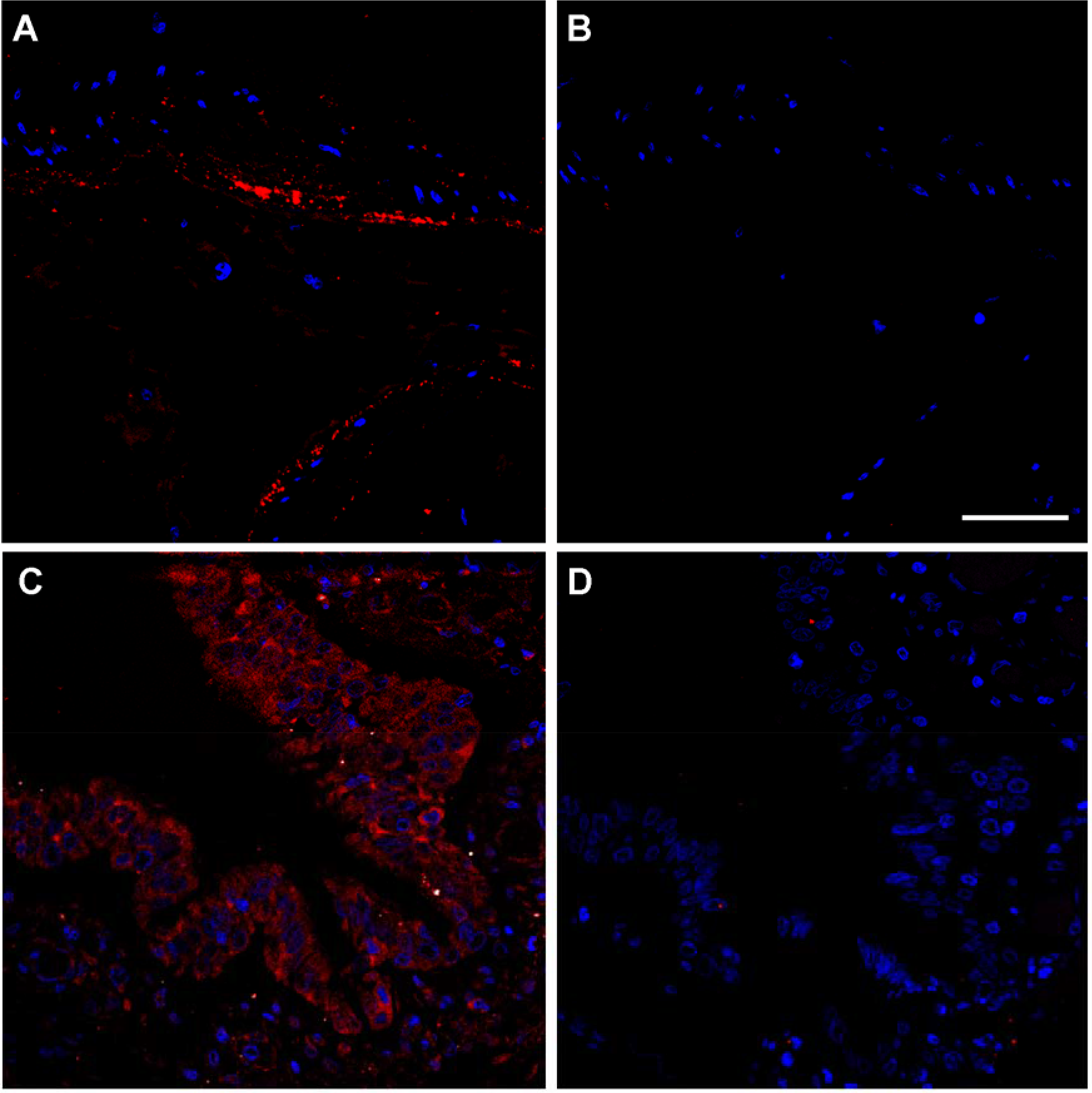
Representative confocal miscroscopy images of lung tissue sections from non-COVID-19 autopsy specimens showing RAMP1 immunoreactivity (red) in the smooth muscle cells of an artery (A,B) and in the bronchiolar epithelium (C,D). Absence of the primary antibody (B,D) was used as a negative control. Cell nuclei were counterstained with DAPI (blue). Scale bar = 50 μm.

As previously reported ^4^, histological analysis of the COVID-19 affected lungs revealed epithelial denudation, thromboemboli, hyaline deposits, bronchopneumonia, and proliferation of type II pneumocytes (Fig. 3B), when compared to lungs from non-COVID-19 cases (Fig. 3A). The pattern of RAMP1 immunoreactivity in COVID-19 patients was similar to the one described in non-COVID-19 slides, labeling smooth muscle cells and airway epithelium (Fig. 3C). Remarkably, the proliferating type II pneumocytes displayed an intense immunoreactivity for RAMP1 (Fig. 3D).

**Figure 3.**
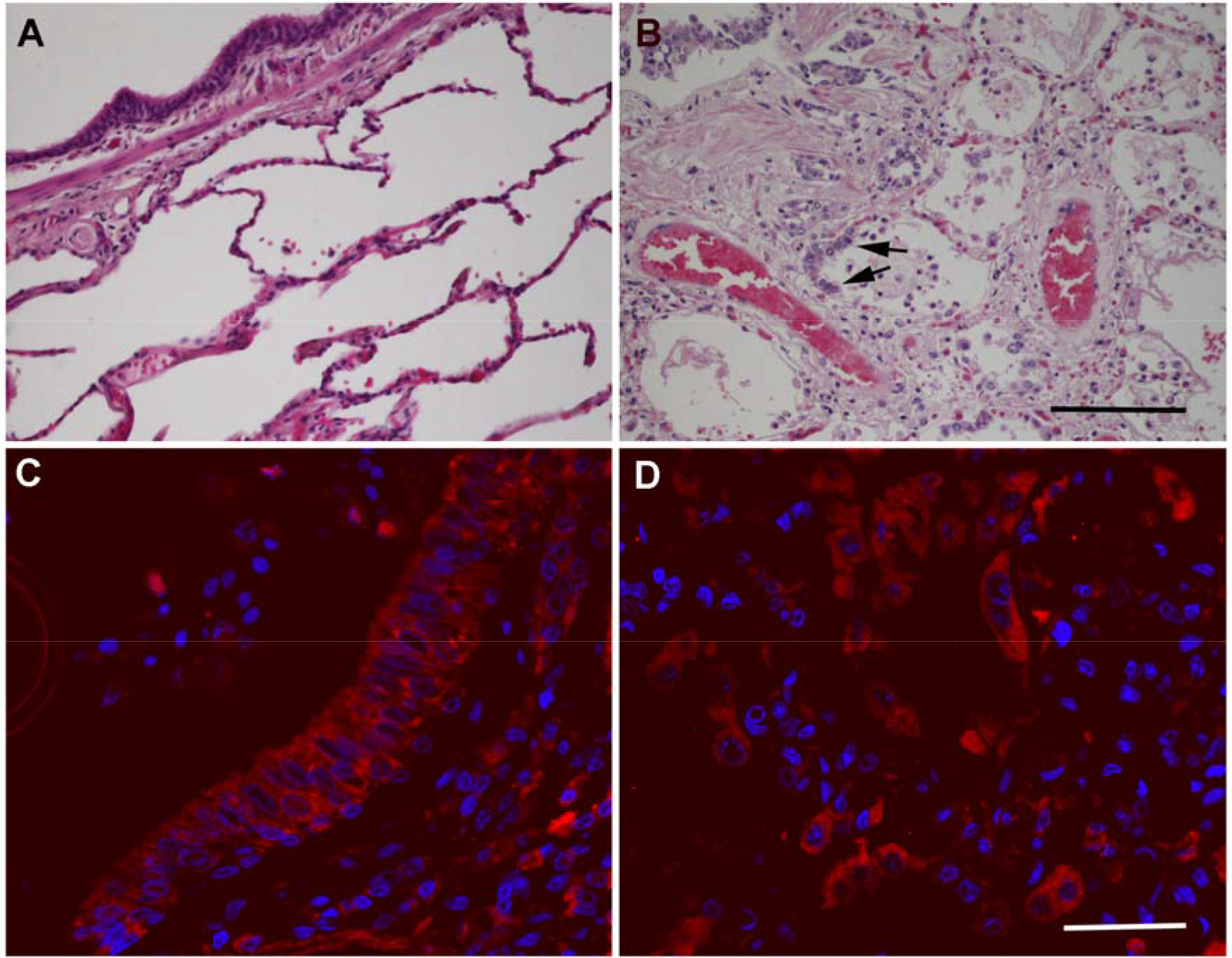
Representative microphotographs of the lung of non-COVID-19 (A) and of SARS-CoV-2-positive samples (B-D). Hematoxylin-eosin stained sections (A,B) show clear histological changes caused by SARS-CoV-2 infection, including proliferation of type II pneumocytes (arrows in B). Immunofluorescence for RAMP1 shows the distribution of this receptor component in the bronchiolar epithelium (C) and the hyperplastic type II pneumocytes (D) in SARS-CoV-2 samples. Scale bar for A and B = 200 μm. Scale bar for C and D = 50 μm.

## Discussion

In this study we have shown that CGRP serum levels become significantly reduced in COVID-19 patients, independently of their disease severity status. In addition, we found that RAMP1 immunoreactivity was predominantly found in the walls of the main vessels and airways, and in the bronchial/bronchiolar epithelium. The hyperplastic type II pneumocytes found in COVID-19 patients were also immunoreactive for RAMP1.

A limitation of the study was that the healthy control group had a lower mean age than the COVID-19 group. This was due to the fact that we wanted to use control samples collected before the pandemia to avoid inclusion of potential asymptomatic subjects, and therefore we had no control over their specific characteristics. Levels of CGRP were not affected by either age or sex in our study, so the age difference should not influence the main conclusions of the study. In addition, the age of the control group is the same as our asymptomatic population, and the CGRP differences between them are remarkable.

A previous commentary suggested that CGRP antagonists may be helpful in the fight against COVID-19 ^24^ but, to the best of our knowledge, this is the first report on the reduced levels of the peptide in patients. CGRP belongs to a peptide family including also calcitonin, adrenomedullin, and amylin ^25^. Interestingly, it has been published that the circulating levels of calcitonin ^26^ and adrenomedullin ^27^ are elevated in COVID-19 patients, probably as a consequence of widespread inflammation and/or blood vessel damage. No data are available for amylin. That CGRP goes in the opposite direction suggests a very specific regulation for this peptide.

The circulating levels of CGRP in patients which received chronic treatment with proton pump inhibitors were lower than in other patients. Although these treatments may slightly confound the final CGRP levels among the COVID-19 patients, we showed that COVID-19 patients not receiving these treatments had still significantly lower levels of the peptide. Even though no previous reports investigated the blood levels of CGRP in patients receiving these drugs, there are some animal models where exposure to omeprazole and other inhibitors significantly reduced the expression of CGRP in dorsal root ganglion neurons in the rat ^28^. It has been shown that treatment with proton pump inhibitors is associated with higher odds of infection and with more severe clinical outcomes in COVID-19 patients ^29, 30^. The causes of these serious negative effects of the acid inhibitors are not presently known, but our results suggest that some of them may be mediated by a decrease in CGRP levels.

Previous studies have established the behavior of CGRP in the presence of viral infections. For instance, respiratory syncytial virus infections decrease CGRP expression in the lung, and treatment with CGRP abolished airway hyperresponsiveness ^31^. This is a behavior very similar to what we have observed in COVID-19 patients. The involvement of CGRP in reducing viral-induced hyperresponsiveness may be very helpful in the treatment of COVID-19 patients. In addition, it has been shown that CGRP may inhibit HIV-1 transmission ^32^. Although the transmission mechanisms are different between these viruses, it might be interesting to test whether CGRP has any impact on SARS-CoV-2 life cycle.

The effects of CGRP as a potent vasodilator are well known ^10^. A significant reduction in the levels of CGRP may result in widespread vasoconstriction in the lung, which may contribute to promoting vascular occlusion, one of the milestones of COVID-19 ^33^. Another critical function of CGRP is its influence on the immune system. SARS-CoV-2 infection stimulates the production of prostaglandin D_2_ which, in turn, upregulates the ILC2 response, leading to sequestration of lymphocytes into the lung and peripheral lymphopenia ^34^. CGRP is an inhibitor of ILC2 ^16^, so we can infer that the lower levels of the peptide observed in COVID-19 patients would participate in worsening immune dysfunction and lymphopenia.

Our immunofluorescence study showed immunoreactivity for RAMP1 in the walls of the major blood vessels, bronchi, and bronchioli, as well as in the epithelium of conducting airways. To the best of our knowledge this is the first morphological description of RAMP1 immunoreactivity in the human lung, but it coincides very closely with the location of CGRP binding sites as studied by radioactive ligand binding ^12^. COVID-19 samples had the same distribution of RAMP1 than non-COVID-19 lungs, with the exception that proliferating type II pneumocytes, which are found in many COVID-19 patients ^4^, were also positive for RAMP1. In contrast, normal type II pneumocytes in non-COVID-19 samples did not express this receptor component. This result may explain the higher expression of RAMP1 that was found in COVID-19 lung samples using multiplex gene expression analysis ^6^. The upregulation of the CGRP receptor may reflect a feed-back mechanism trying to compensate for the reduction in systemic CGRP levels. Also, the high expression of RAMP1 in proliferating type II pneumocytes may indicate an important implication of CGRP in the repair of the respiratory epithelium subsequent to damage caused by viral infection ^35^.

All these evidences suggest that restoring CGRP levels may constitute a new intervention for COVID-19 patients. Although CGRP inhibitors have been more developed pharmacologically, due to their application in fighting migraines ^36^, several experimental strategies have been proposed for elevating CGRP levels, with positive outcomes ^37-39^. Application of these CGRP “donors” to COVID-19 patients may improve several critical manifestations of the disease, such as proper angiogenesis ^40^, vasodilatation, bronchial epithelium repair, and immune system regulation.

## Data Availability

Complete datasets will be provided to any researcher

